# Features importance in seizure classification using scalp EEG reduced to single timeseries

**DOI:** 10.1101/2021.07.28.21261310

**Authors:** Sébastien Naze, Jianbin Tang, James R. Kozloski, Stefan Harrer

## Abstract

Seizure detection and seizure-type classification are best performed using intra-cranial or full-scalp electroencephalogram (EEG). In embedded wearable systems however, recordings from only a few electrodes are available, reducing the spatial resolution of the signals to a handful of timeseries at most. Taking this constraint into account, we tested the performance of multiple classifiers using a subset of the EEG recordings by selecting a single trace from the montage or performing a dimensionality reduction over each hemispherical space. Our results support that Random Forest (RF) classifiers lead most efficient and stable classification performances over Support Vector Machines (SVM). Interestingly, tracking the feature importances using permutation tests reveals that classical EEG spectrum power bands display different rankings across the classifiers: low frequencies (delta, theta) are most important for SVMs while higher frequencies (alpha, gamma) are more relevant for RF and Decision Trees. We reach up to 94.3% ∓ 5.3% accuracy in classifying absence from tonic-clonic seizures using state-of-art sampling methods for unbalanced datasets and leave-patients-out fold cross-validation policy.

## I. Introduction

Epilepsy manifests through seizures which occurs uncontrollably [1]. Several types of seizures exist based on semiology, symptomatic experience and electrophysiological signatures [2]. Patients with epilepsy can display several seizure types [3], and the monitoring of seizures for forecasting and detection is a subject of intense research [4], [5]. Recent advances in the field use elaborate methods from machine learning to analyse EEG timeseries and automatically extract the most relevant features from the signals to perform the detection or classification task [6], [7].

An issue of those deep learning methods lies in the lack of interpretability of the abstract features learned by the deep neural network to perform the task [8]. Using manually engineered features can help for interpretation but typically these perform suboptimally on electrophysiological recordings [9], [10], therefore highlighting a tradeoff challenge between efficiency and interpretability.

Another challenge in patient monitoring is the movement away from the hospital settings and towards recording spontaneous seizures from a wearable device at home or in daily life [11], [12]. A major drawback of these wearable recordings is that their spatiotemporal resolution is further constrained in order for the device to be minimally inconvenient for the patient [13], [14].

Here, we confront those two challenges by reducing the spatiotemporal resolution of EEG signals to single timeseries per hemisphere and training classifiers on these series using engineered features. A combination of preprocessing methods, sampling algorithms and classifier types is explored systematically. The importance of each feature for each classifier is assessed using the best combination of preprocessing steps.

## II. Materials & Methods

### A. Dataset

We used the EEG Corpus from the Temple University Hospital (TUH) dataset [15]. The data was recorded using scalp EEG with 20 electrodes following the standard international 10-20 system, and timeseries were analyzed using the longitudinal transverse bipolar montage. All patient recordings which sampling frequency were different from 256 Hz were resampled at 256 Hz prior to preprocessing.

### B. Pre-processing and feature extraction

Samples of preictal and ictal periods were created using 4s sample size (as in [16]). Periods shorter than the sample size were discarded. A combination of several preprocessing steps were performed on each sample:

#### Transformation to single timeseries per hemisphere

the signals from electrodes of each hemisphere were reduced to a single timeseries by either taking *(a)* the average over electrodes from the right and left hemisphere separately; *(b)* the strongest vector of the principal component analysis over the left and right hemisphere separately; or *(c)* the difference between electrodes (F7-T3 and F8-T4 for left and right hemispheres, respectively).

#### Normalization

the signal was then normalized for each sample using a z-score normalization over the sample size, i.e. 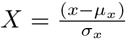, and further zero-centered using the moving-average over a 1s sliding window.

Fig. 1 illustrates the preprocessing of EEG samples. Features are computed as the average power over the alpha (*α*, 8-12 Hz), beta (*β*, 12-25 Hz), gamma (*γ*, 25-80 Hz), delta (*δ*, 0-4 Hz) and theta (*θ*, 4-8 Hz) frequency bands.

**Fig. 1.**
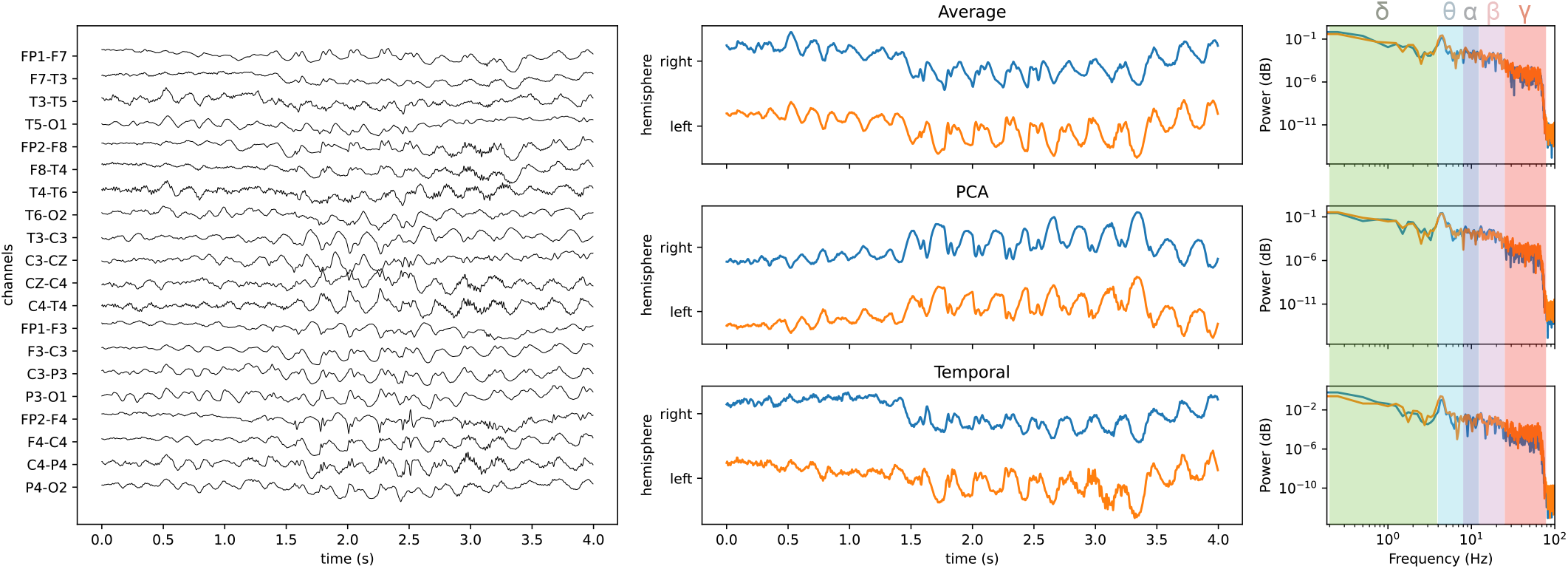
Example of a 4s EEG sample (left) and its reduction to one timeserie per hemisphere (middle) using averaging over channels, PCA, or a subset of temporal channels. Power spectral features are used for *α, β, γ, δ* and *θ* bands (right).

### C. Sampling of imbalanced classes

Class imbalance can lead to skewed classification accuracies towards the class with most samples [17]. We alleviate this problem by sampling from the classes during training and testing phases using several methods from the `imblearn` library [18]:

- *Random Under Sampling* is a method that picks random samples (without duplicates) from the majority class(es) until the number of samples equals the minority class. This method is therefore referred to as unbiased from the sample distribution.
- *Cluster Centroids* uses the samples from the majority class that are closest to the class center determined by K-mean clustering. This methods is therefore biased towards samples that represent the average of the class.
- *Near-Miss* under-sampling is a method that takes the samples from the majority class which are on average the closest to the samples from the minority class [19]. This method makes the classification more difficult as samples are selected to be least discriminable between classes.

### D. Classification

We assessed several types of classifiers to discriminate between absence and tonic-clonic seizures. Here, we briefly summarize the different characteristics of each classifier:

- *Support Vector Machines (SVMs)* try to find the maximally separating hyperplanes between samples from 2 classes (and do so repeatedly for each combination of classes in multi-class classification). The kernel is a similarity function that scores the distance between samples in feature space. We assess the score of SVMs using a linear kernel (simplest case) and a non-linear radial-basis function (RBF) kernel. Since our feature space is much smaller than our number of samples, the method was not prone to over-fitting and the regularization parameter was set to *C* = 1.
- *Decision trees* are hierarchical structures (connected nodes) whereby each branching represents conditions over features which aggregate through the depth of the tree to define class labels. The conditions for the split criterion are found by maximizing the entropy at each node of the tree. We set the maximum depth of the tree to be the number of features (5) and a minimum of 2 samples is necessary to split a node into branches.
- *Random Forests* are a set of many decision trees which outputs are combined to form the best weighted conglomerate. Individual trees are created at random based on feature values at the beginning and then refined through training.

### E. Cross-validation

We performed our analysis using 3-fold cross validation scheme with leave-patient-out policy. This means that for each split, all samples from a patient are either in the training set or the test set, without overlap. This prevents over-fitting on the specific patient trace which is commonly observed in standard n-fold cross validation policies ([6], [7]). We used the Stratified Group K-fold splitting module from `scikit-learn` [20], whereby classes are seizure labels and groups are individual patients.

### F. Feature importance

The importance of each of the spectral band features to the overall performance of trained classifiers is assessed through a random permutation test over samples (rows), one feature (column) at a time. For each permutation, the classifier is re-trained using the permuted feature values and the score is compared to the originally trained classifier’s score [21]. The score’s difference is averaged over 100 permutations to give a value between 0 (low importance) to 1 (high importance), and was performed using the `scikit-learn` library [20].

## III. Results

We created a pipeline combining sample creation from raw EEG, preprocessing, feature extraction, dataset splitting for 3-fold cross validation, sampling, classification and extraction of feature importances. We first assessed the performance of each classifier using a combination of preprocessing steps and data samplers. We then analyse which features play a major role in discriminating between seizure types.

### A. Performance of the different classifiers across preprocessing methods

Figure 2 shows the performance of the 4 classifiers using the 3 samplers and 3 different dimensionality reduction techniques at preprocessing. We observe that performing a PCA over the EEG instead of using a subset of temporal electrodes systematically increases the accuracy of the downstream classification (mean +17.4%, SD 8.8%). It is also observed that the PCA does better than averaging across all electrodes per hemisphere (mean +0.9%, SD 3.7%).

**Fig. 2.**
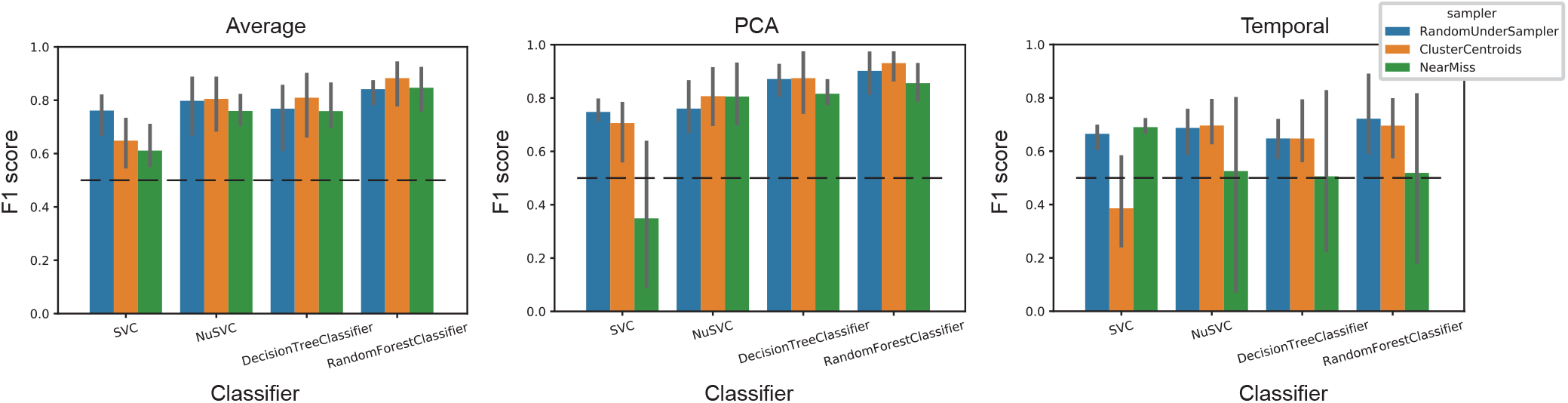
Overall accuracy of each classification scheme computed by F1-score across preprocessing methods. The horizontal dashed line is the score under the null hypothesis, i.e. when the class is predicted randomly.

The difference in sampling method accounts to 9.9% (SD 2.8%) of classification performance. Cluster centroids resulted in the highest performances. Surprisingly, a near miss sampling can outperform a random sampling (or perform similarly) when using a dimensionality reduction technique such as PCA or averaging. Also, linear SVM is more sensitive to the sampling methods that non-linear SVM, decision tree and random forest classifiers.

### B. Feature importance differs across classifiers

Lastly, we assess the contribution of each feature to the classification accuracy. This is performed by randomly permuting row entries of a feature column and re-training the classifier to assess its new accuracy using the permuted feature values. The drop in accuracy is indicative of the feature importance of the column being shuffled. Since preprocessing using PCA and cluster centroids sampling reached the best classification accuracy, we show feature importances for each classifiers using this preprocessing methods but similar results were observed using averaging across electrodes and other sampling methods.

Figure 3 shows that the feature importance for the delta frequency band (0-4 Hz) exceeds all others for SVM classifiers, while higher frequency bands (alpha and gamma) play the major role for decision tree and random forest classifiers. This indicates that higher frequencies in the EEG play a subtle but essential role in improving the classification performances across seizure types and across the classification methods that we explored.

**Fig. 3.**
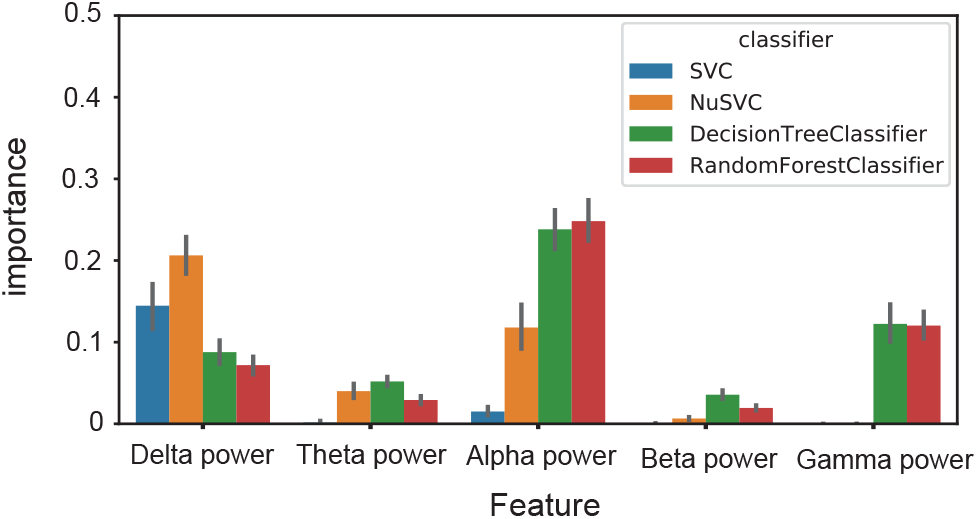
Feature importance across classifiers.

## IV. Conclusion

We assessed several pre-processing methods in the context of EEG signal classification between tonic-clonic and absence seizures. By systematically comparing classification performances across timeseries normalization schemes, sampling from imbalanced classes and dimensionality reduction techniques, our results demonstrate that applying a PCA over the whole EEG signals leads to better outcomes than using only a subset of electrodes or averaging across electrodes. This indicates that sacrificing temporal precision for spatial integration of the signals across the scalp is beneficial for this seizure type classification task. It is clear that in the designing of wearable systems for patients monitoring using EEG, recordings from many electrode spatially distributed over the scalp gives better classification outcomes than using a single electrode. This is especially important for patients experiencing a wide range of seizure types or when the seizure semiology evolves across the span of the disorder (e.g. transitioning from absence to tonic-clonic [22]).

Our work also paves the route for more interpretable results of machine learning outputs. This is especially important for medical applications, since a mechanistic understanding of the AI systems can permit better comprehension of the processes at play in patients. Others have reviewed features of interest for seizure detection and classification [10], [23]. Our results indicate that while low frequency EEG component (i.e. delta band) is most relevant for classification using SVM, more elaborate classifiers devote greater importance to the higher frequencies (alpha and gamma bands). Since the interplay of low and high frequency discharges during seizure is complex and specific to certain seizure types, our results suggest that each classifier type is picking up on those features differently. A next step will involve modeling of seizure dynamics [24], and optimization of model parameters to reproduce the different classes synthetically as conceptually introduced in a previous study on classifying transcranial magnetic stimulation responses [9].

## Data Availability

This study used the openly accessible dataset from Temple University Hospital EEG Corpus.

https://www.isip.piconepress.com/projects/tuh_eeg/

## V. Acknowledgments

We are indebted to Joseph Picone, Iyad Obeid and their team of researchers, clinicians, engineers and technicians at Temple University who collected and curated the dataset.

